# “The drug development pipeline for glioblastoma - a cross sectional assessment of the FDA Orphan Drug Product designation database”

**DOI:** 10.1101/2021.01.01.21249120

**Authors:** Pascal Johann, Dominic Lenz, Markus Ries

## Abstract

**Background:** Glioblastoma multiforme (GBM) is the most common malignant brain tumor among adult patients and represents an almost universally fatal disease. Novel therapies for GBM are being developed under the orphan drug legislation and the knowledge on the molecular makeup of this disease has been increasing rapidly. However, the clinical outcomes in GBM patients with currently available therapies are still dismal. An insight into the current drug development pipeline for GBM is therefore of particular interest.

**Objectives:** To provide a quantitative clinical-regulatory insight into the status of FDA orphan drug designations for compounds intended to treat GBM.

**Methods:** Quantitative cross-sectional analysis of the U.S. Food and Drug Administration Orphan Drug Product database between 1983 and 2020. STROBE criteria were respected.

**Results:** Four orphan drugs out of 161 (2,4%) orphan drug designations were approved for the treatment for GBM by the FDA between 1983 and 2020. Fourteen orphan drug designations were subsequently withdrawn for unknown reasons. The number of orphan drug designations per year shows a growing trend. In the last decade, the therapeutic mechanism of action of designated compounds intended to treat glioblastoma shifted from cytotoxic drugs (median year of designation 2008) to immunotherapeutic approaches and small molecules (median year of designation 2014 and 2015 respectively) suggesting an increased focus on precision in the therapeutic mechanism of action for compounds the development pipeline.

**Conclusion:** Despite the fact that current pharmacological treatment options in GBM are sparse, the drug development pipeline is steadily growing. In particular, the surge of designated immunotherapies detected in the last years raises the hope that elaborate combination possibilities between classical therapeutic backbones (radiotherapy) and novel, currently experimental therapeutics may help to provide better therapies for this deadly disease in the future.

**Article summary:** *Strengths and limitations:*

- This study provides a quantitative overview on the drug development pipeline for pediatric and adult oncology in general and specifically for the indication glioblastoma
- Analyzing the therapeutic mechanisms of designated compounds in glioblastoma reveals an increased focus on personalized and targeted therapies
- The precise reasons for failure of approved drugs and for withdrawal of approved drugs in glioblastoms are unknown
- For the analysis, only the databases “FDALabel” (https://nctr-crs.fda.gov/fdalabel/ui/search) and the FDA Orphan drug product designation database (https://www.accessdata.fda.gov/scripts/opdlisting/oopd/) were considered

## Introduction

High grade gliomas account for the majority of brain tumor related deaths in children and adults. Considering all age groups together, glioblastoma multiforme represents the most common malignant brain tumor (43,5% of all malignant brain tumors [1]).

Albeit being rare in absolute numbers, glioblastomas represent a universally fatal disease class for approximately 15,000 patients per year in the United States [1]. While the last years have seen a surge in publications that highlight intertumoral and intratumoral diversity [2, 3] in these tumors, our growing understanding of the pathophysiological processes that underlie the disease could so far not yet be translated into therapeutic success.

By now, a wealth of studies has identified the typical genetic alterations that occur in glioblastoma: Mutations in *IDH1* or (in pediatric glioblastomas) the two frequently occurring histone *H3*.*3* gene mutations (*H3*.*3:* pK27M and *H3*.*3*: pG34R/V) are just two examples of common genetic lesions which define distinct molecular classes of glioblastoma. The well-known genetic lesions identified in glioblastoma have subsequently lead to the identification of epigenetic and transcriptomic mechanisms which perpetuate the disease: examples for this are the hypermethylation of CpG islands in IDH1-mutant glioblastoma [4] or the loss of histone H3.3 K27me3 in H3.3 mutant glioblastomas [5].

Despite the vast increase in knowledge on genome, epigenome and transcriptome of glioblastoma, clinical outcomes have not changed and drug development in glioblastoma is lagging behind the significant advances in glioblastoma (epi)genomics. While some of these genetic targets can be used therapeutically, the majority of them is unsuitable as a drug target but may offer the prospect to be used in immunotherapy.

Thus, there is an unequivocal medical need for novel compounds or combinations of compounds that are able to put a hold on disease progression.

The U.S. Orphan Drug Act of 1983 was intended to incentivize drug development in rare diseases including rare cancers by providing protocol assistance, orphan grants programs, tax credit for 50% of clinical trial costs, U.S. Food and Drug Administration (FDA) fee waiver, and 7 years of marketing exclusivity [6]. Between 1983 and 2015, more than a third of all orphan drug approvals (N= 177 out of a total of 492, i.e., 36%) were related compounds intended to treat rare cancers [7].

While there may be manifold reasons for a clinical failure of novel drugs, a comprehensive view on the status of designated compounds for the indication glioblastoma is still lacking. In particular, it remains unclear which substance classes and therapeutic principles for glioblastoma have entered the market or are under development. This knowledge is instructive as the pharmacological principles which underlie the designated drugs may have changed over time and thus may mirror the different directions of brain tumor research. We aim at analyzing lessons which we have learned by assessing successes and failures in orphan drug development in glioblastoma. Therefore, we present a cross-sectional, quantitative clinical-regulatory insight into the status of FDA orphan drug designations for compounds intended to treat GBM. This study covers the period between January 1983 and August 2020.

## Methods

STROBE criteria (Supplemental table 1) were respected for planning, conduct, analysis, and reporting of this study [8]. We accessed the Orphan Drug Product designation database on 30.07.2020 at https://www.accessdata.fda.gov/scripts/opdlisting/oopd/ and downloaded the information on all designated drugs. The list of drugs was then manually cleared from non-oncological indications. An allocation to the field of “pediatric oncology” or “adult oncology” was performed by a board certified pediatric oncologist.

Disease entities which typically occur both in adult age and in the field of pediatric oncology (such as lymphomas and osteosarcoma for instance) were allocated to both categories. Others which almost typically occur in pediatric oncology such as ALL were considered only for this area.

Subsequently, designated drugs which were intended to treat glioblastoma were characterized according to their mode of action in the pharmacological classes “cytostatics”, “cellular product-virus” or “targeted therapies”. Targeted therapies were defined as substances for which at least one molecular target could be identified by literature research [9]. Compounds which could not be classified unequivocally were categorized as “others” – this class also contained compounds that are being used as diagnostics.

In order to independently verify whether there were approved drugs for the treatment of glioblastoma that were not listed in the U.S. Food and Drug Administration Orphan Drug Product database, we conducted a full text search in the FDA drug label database (FDALabel, https://nctr-crs.fda.gov/fdalabel/ui/search). Search terms were “glioblastoma” in the section “indications and usage”. The database was accessed on 28 October 2020.

Findings were juxtaposed to the approved compounds identified from the search in Orphan Drug Product designation database as described above. In addition, we cross-validated whether or not the compounds identified from the search in FDALabel were registered as orphan drugs.

Standard methods of descriptive statistics were applied. In particular, continuous variables were summarized with mean, standard deviation, and median, minimum, and maximum values whereas categorical variables were summarized with frequencies and percentages. Analyzed groups included 1) approved and 2) designated compounds intended to treat glioblastoma.

In order to determine the number of approved drugs, we first queried the downloaded data from the Food and Drug Administration Orphan Drug Product database for FDA approved drugs and the curated the list by eliminating duplicate terms. Likewise, the data were analyzed for designated compounds. In addition, we analyzed number and characteristics of orphan drug designations for glioblastoma that were subsequently withdrawn. In order to put our findings on orphan drug designations for glioblastoma into perspective within a global oncological context we analysed orphan drug designations for all oncological indications currently listed in the US Food and Drug Administration Orphan Drug Product database.

For statistical analysis and graphical display we used the software R (version 3.5.0). For plots, the program library ggplot2 was employed. We used the CONSORT checklist when writing our report [10]

## Results

### Approved drugs for glioblastoma

A total of four compounds (Table 1) were approved by the FDA for the indication glioblastoma. Three of them were therapeutic compounds, one is 5-aminolevulinic acid which is a photo-diagnostic substance for the intraoperative detection of resection margins [11].

These results were in line with the findings of the FDALabel database query: bevacizumab, carmustine, and temozolomide list the indication “glioblastoma” on their FDA approved labels. All of these drugs have been used in the clinical setting, however with daunting results and no improvement in the outcome of glioblastoma [12].

The small number of approved compounds prompted us to further explore the drug developmental landscape in this disease.

*Table 1: An overview on approved drugs for the indication “Glioblastoma”*

### Designated drugs in glioblastoma

Given the low number of approved drugs that can be used in the clinical setting, we next sought to get an overview on the drug development landscape in glioblastoma, trying to quantify and qualify the drugs which are in the pipeline for this indication

Overall, 162 compounds had an orphan drug designation for glioblastoma. We found a steady increase of orphan drug designations over time (median per year 4)-only from 2016 on, their number was decreasing. To be able to put these findings in GBM into a global oncological drug development context, we assessed the spectrum of all oncological - and pediatric oncological diseases with with orphan drug designations.

Our analysis here yielded 4618 compounds (out of a total of 5513 orphan drug designations for any rare disease= 83%, Figure 2 A) which were designated for either a pediatric or an adult oncological entity. As in glioblastoma, since 1984, the number of orphan drug designations for adult oncology per year rose continuously with a peak of 342 compounds in 2016. As expected, the number of compounds targeting typical disease entities from adult oncology was consistently higher than in pediatric oncology (on average 37 % higher). The numeric pattern over time appeared to be similar in pediatric and adult orphan drug designations.

**Figure 1:**
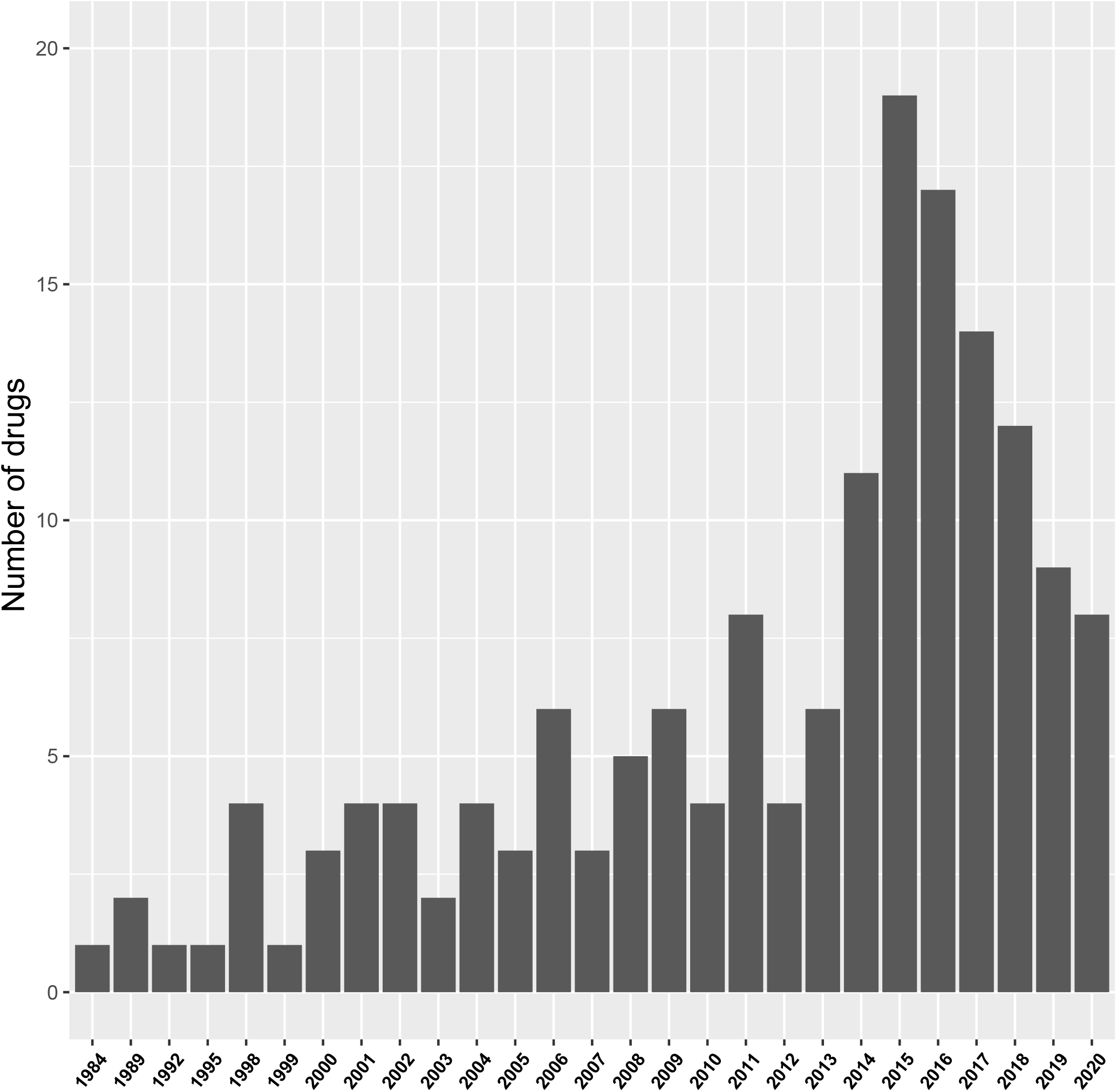
Barplot shows the number of orphan drug designations for the indication glioblastoma per year

**Figure 2.**
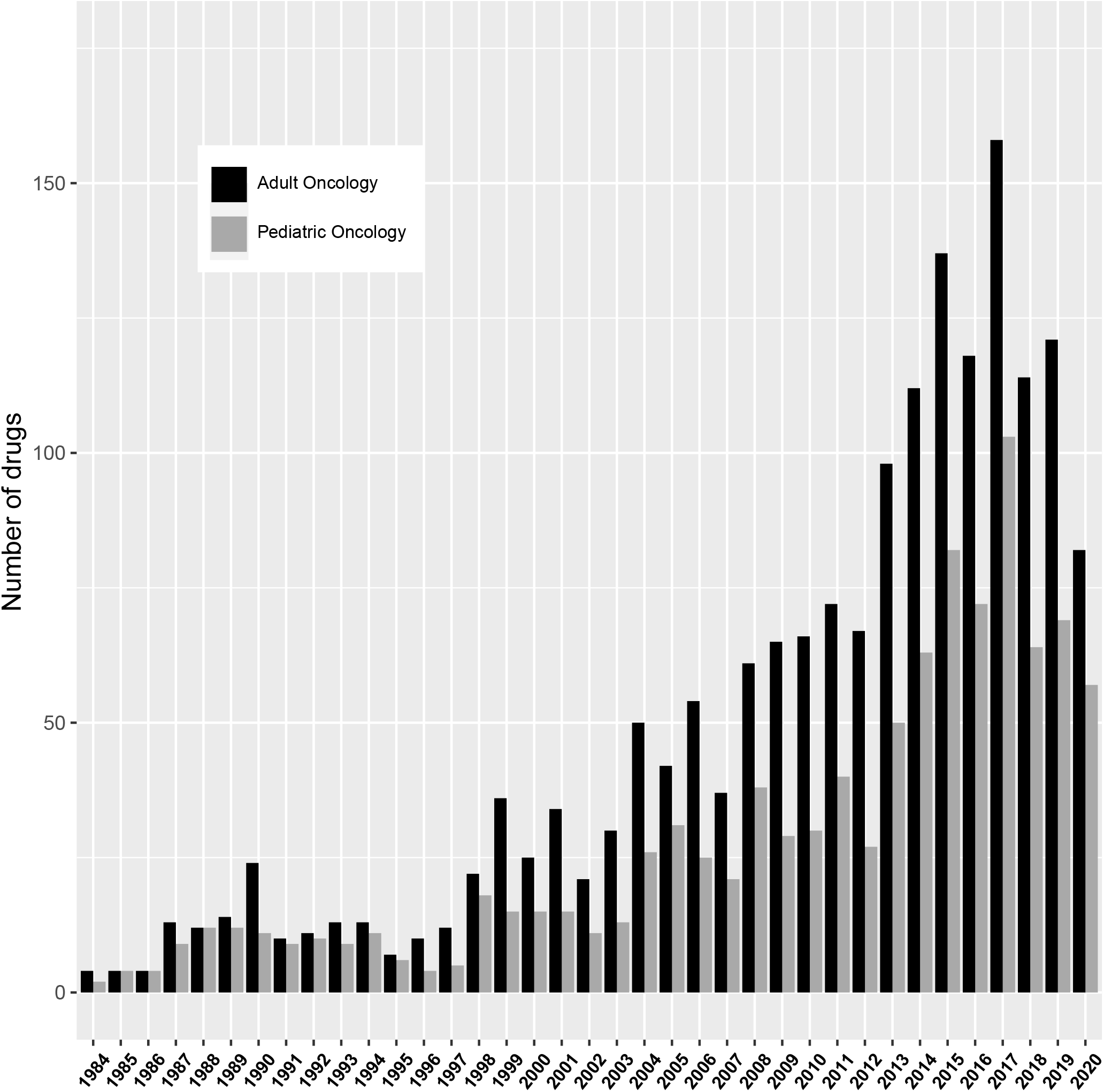
Barplot shows the number of orphan drug designations in pediatric oncology and in adult oncology per year.

To better understand the intended indications of these designated compounds, we then analyzed, which tumor entities are targeted by these drugs.

Figure 3 shows the frequency distribution of FDA orphan drug designations for their respective oncological indications. Most oncological orphan drug designations for the adult patient population were granted for lymphoma, pancreatic cancer, and glioblastoma. In contrast, lymphomas, glioblastoma and AML received the majority of orphan drug designations for pediatric cancers.

**Figure 3.**
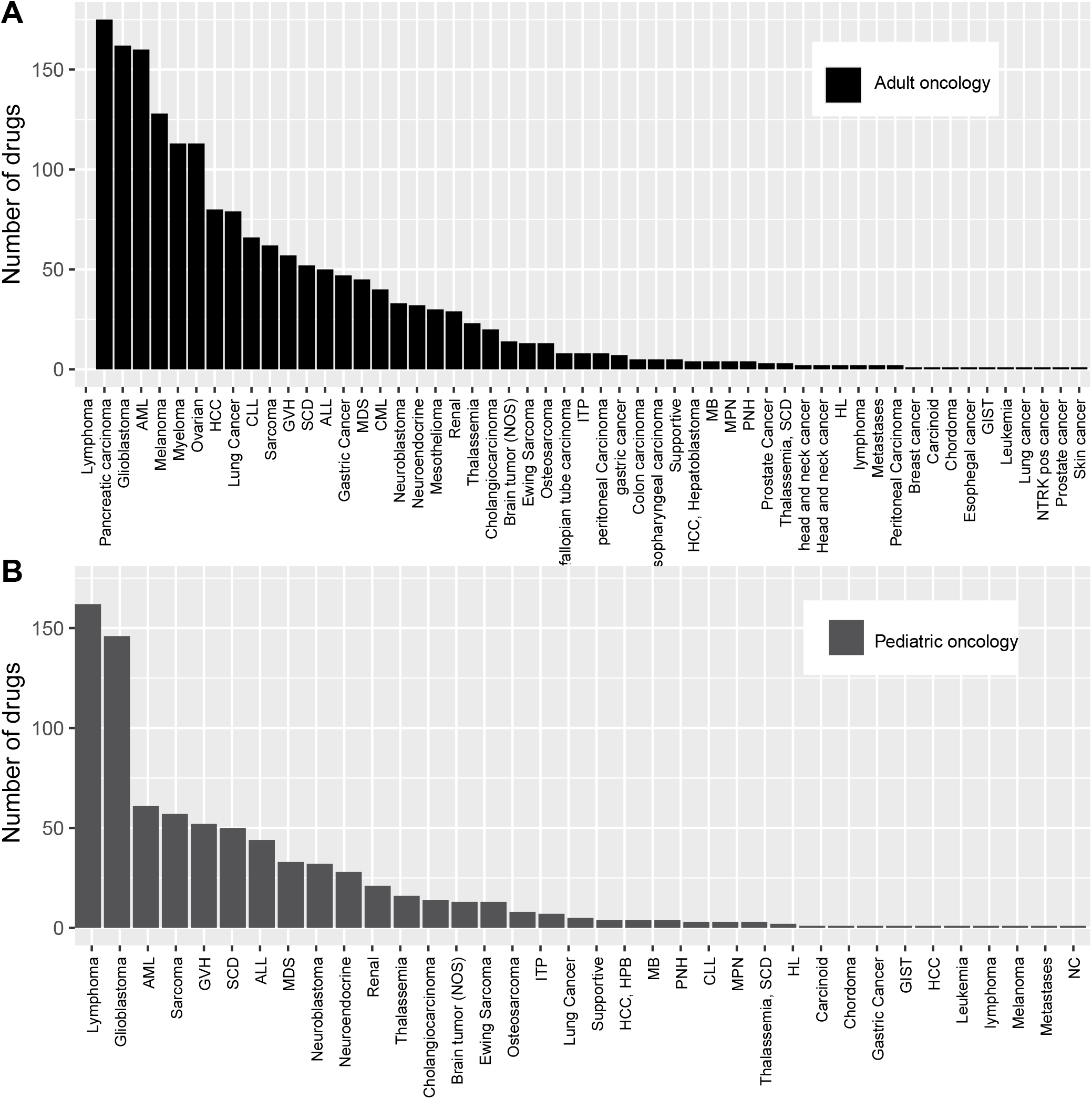
Barplot shows the distribution of entities in pediatric and oncological entities with orphan drug designations.

(Figure 3 A, B). Although some of these categories are quite heterogeneous and comprise different entities (such as lymphomas which are in fact a group of genetically heterogeneous diseases associated with divergent outcome), the predominance of these groups is remarkable as they do not represent the oncological indications which occur most frequently but which are associated with a high mortality. Thus, the designated compounds in fact address the unmet medical need of cancers which are associated with a high mortality despite not being the most frequent ones [13].

### Withdrawn orphan drug designations in glioblastoma (Table 2)

Studying the compounds designated for glioblastoma which were subsequently withdrawn from the market is instructive as it may highlight potential mechanistically interesting substances that never reached the clinic:

Our search in the FDA approved drugs database revealed 14 designated compounds which were subsequently withdrawn from the drug development pipeline. Some of these drugs were classical cytostatics, others displayed more innovative modes of action: cilengitide, an integrin inhibitor, with a well characterized mode of action [14] and substantial preclinical evidence was among the withdrawn substances. Other, less well-known substances included the glutamate receptor inhibitor talampanel and cintredekin besudotox, an IL13 conjugated toxin, specifically targeting glioblastoma. Reasons for these withdrawals were not published and are therefore, unfortunately, not known.

*Table 2 gives an overview on withdrawn drugs that were approved for the indication glioblastoma*

### Pharmacological classes of designated drugs in glioblastoma

We next characterized the pharmacological classes which were designated per year: we therefore allocated the designated compounds into the broad categories “cytostatics”, “targeted therapy”, “Cellular product/ Virus” and “others”. The latter constitutes a heterogeneous group of substances comprising intraoperative fluroescent dyes (as diagnostics), peptide vaccinations or repurposed drugs such as canabinoids which are approved for other indications and were subsequently found to display anti-neoplastic properties (“repurposing”).

When regarding the designation per compound class over time, we found that the median year of designation for cytostatic drugs was 2008 (Figure 3). With an increasing knowledge on both the genetic makeup of glioblastomas and the resulting therapeutic targets, the years 2010-2020 saw an increase in small molecule inhibitors, directed against specific molecular structures (Figure 4 A). An investigation on the nature of these therapeutic targets revealed a high diversity: Compounds directed against VEGF or the VEGFR were most frequently found, but there were also molecules directed against EGFR – a molecule prototypically mutated in subsets of adult glioblastoma [2].

**Figure 4.**
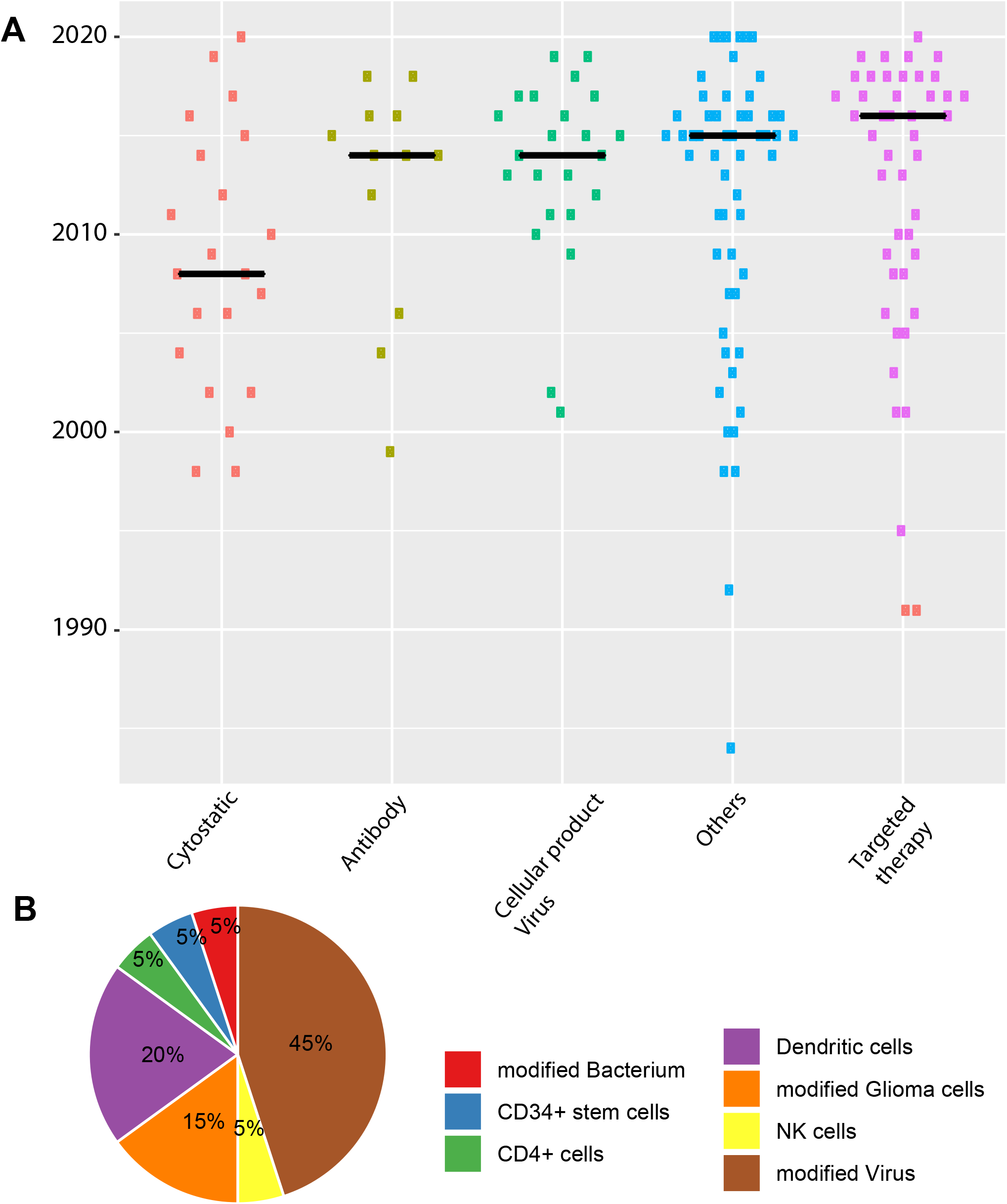
A Boxplot shows the substance classes of designated drugs in GBM, B Pie chart shows the mode of action of designated immunotherapies/ cellular therapies for glioblastomas.

Similarly, immunotherapeutic approaches including dendritic cell vaccinations, or NK cell/T-cell based therapies represents a focus of the last years compound designations. As cellular and viral therapies represent a very diverse group, we dissected this category further (Figure 4 B): Remarkably, 45% (9/20) of all therapeutics proved to be virus based, the majority of which being oncoloytic viruses. The dendritic cell based therapies, the second largest group, were mostly stimulated with autologous tumor lysates or with synthetic peptides derived from glioblastomas, aiming to elicit an anti-tumor immune response in the host.

Although none of the latter therapeutics has been approved for glioblastoma so far, the number of designated drugs in this category points to a high potential of these compounds in the clinic.

## Discussion

### Approved drugs in glioblastoma

The overall increase in orphan drug designations for the whole oncological field is also seen when only regarding glioblastoma (with an average of six designated drugs per year). In stark contrast to the number of designations, only six compounds were approved for this entity in the last 30 years – the most recent substance being bevacizumab, an antibody that targets VEGF, which however did not show a survival benefit in large, placebo-controlled studies[15].

Other approved compounds for GBM include cytostatic drugs such as carmustine or temozolomid. Temozolomid has become a frequently used standard therapy due to its generally favourable toxicity profile. It is one of few drugs for which a biomarker has been identified: The MGMT promoter governs expression of the corresponding gene. It represents the most important resistance mechanism to an alkylating therapy and its hypermethylation has been associated with a better outcome [16].

It is remarkable that so far no cellular or virus based immunotherapy has been granted approval by the FDA, although there have been promising preclinical and clinical [17] studies suggesting a potential benefit.

Of particular during the last years, the number of designated cellular therapies has increased. Some of them employ T-cells with chimeric antigen receptors, others back on dendritic cell vaccinations. It might be assumed that some of these innovative therapeutics will eventually make it to approval for GBM.

### Spectrum of indications for designated drugs

Overall, our study revealed a wealth of compounds designated for oncological indications and an increase in drug designations per year over time. It is notable, that the majority of designated compounds in adult oncology targeted lymphomas, pancreatic cancer and glioblastoma. This does not necessarily reflect the epidemiological spectrum of malignant diseases with breast cancer, lung cancer and prostate cancer being the most frequent cancers. When, however, considering cancer mortality from these entities, the designations certainly do meet a medical need.

### Withdrawn drugs in Glioblastoma

Several orphan drug designations for glioblastoma were subsequently withdrawn without ever having been approved. Unfortunately, the precise reason why is not known. It would be interesting to capture this information transparently in public or even in the clinicaltrials.gov database as this would allow the scientific community to learn from previous experiences, and potentially avoid unnecessary exposure of subjects to clinical research. Possible reasons for failure may include a flawed scientific rationale, flawed trial design, lack of funding, unprecedented regulatory environment, or unsustainable business (https://termeerfoundation.org/collaborations accessed 06 October 2020 [18]).

### Drug safety considerations and innovative aspects of drug development in glioblastoma

Usually orphan drug development programs involve less patients and less clinical trials than non-orphan drug development programs. No approved drug for glioblastoma was withdrawn. This indicates that there were no major safety issues in the orphan drug development process in this area that were detected in the post-approval pharmacovigilance process. With respect to innovation in the process of granting approval to novel drugs, there is certainly room to expedite this process: The only targeted drug among the approved compounds remains bevacizumab. Until today, the impact of the US orphan drug act on the drug development for glioblastoma has been limited: There are only four FDA orphan approvals for the treatment of glioblastoma – one of them (5-aminolevulinic acid.) is a diagnostic compound. There is hope for progress. It is possible that more compounds may successfully reach the clinic, because 60 orphan drug designations have been granted with an increasing tendency in the last 5 years.

### Barriers to a successful translation of preclinical findings to the clinic

Glioblastoma represents a genetically highly complex disease: When progressing, these tumors undergo a complex, molecular evolution which results in an increase of genetic aberrations. Thus, therapies which target only one specific molecular lesion fall short in controlling the manifold other pathways which may contribute to tumor growth. However even combination therapies (including small molecule inhibitors and classical cytotoxic compounds) did not show the desired effects in clinical studies).

Further problems which are encountered in the management of glioblastoma include its invasive nature and the impossibility to achieve a gross total resection due to the infiltrative growth, its high proliferative rate and – associated with the latter-the speed by which resistance mechanisms toward applied therapy emerge. Despite a growing number of designated compounds and molecular directed therapeutics only few of them address the molecular characteristics of the genetic and epigenetic glioblastoma subgroups. As an example, the molecular mechanism that drives H3K27M mutant glioblastomas is well described by now: A sequestration of the enzyme PRC2 [5] leads to a global loss of the repressive histone mark H3K27me3. An inhibitor of enzymes (GSK-J4) that catalyze the demethylation of H3K27me3, thus restoring this mark, has been tested [19] but so far has not reached the status as a designated drug.

There is hope that immunotherapy-either antibody based on the inhibition of PD(L)1 or by using cellular products such as CAR-T cells [20] or dendritic cells - may ultimately improve the outcome for patients with glioblastoma. At the least, these agents represent combination partners that may expand the portfolio of classically used cytotoxic drugs.

### Limitations

This analysis of orphan drug development in the field of oncology is limited to the data provided in U.S. Food and Drug Administration Orphan Drug Product database. Other geographic regions were not included in this study because the FDA database is considered comprehensive. As orphan drug development is generally a global endeavour, we consider the results of this analysis to be generalizable within the context of these limitations.

In summary, we conclude despite the fact that current pharmacological treatment options in GBM are sparse, the drug development pipeline is steadily growing [21].In particular, the surge of designated immunotherapies during the last years raises the hope that elaborate combination therapies between classical therapeutic backbones (radiotherapy) and these novel, currently experimental interventions may help to provide better treatment options for this deadly disease in the future.

Contributors PDJ, MR contributed to the conceptualisation of the project. PDJ, DL and MR performed data collection efforts. PDJ, MR contributed to data analysis, interpretation and writing of the manuscript. All authors contributed to the revision and approved the final version of the manuscript.

## Data Availability

All data relevant to the study are included in the article or uploaded as supplementary information. All data relevant to the study are included in the article.

## Patient consent for publication

Not required.

## Data availability statement

All data relevant to the study are included in the article or uploaded as supplementary information. All data relevant to the study are included in the article.

## Abbreviations

ALL: Acute lymphoblastic leukemia
AML: Acute myeloid leukemia
CAR: Chimeric antigen receptor
CLL: Chronic lymphocytic leukemia
EGFR: Epidermal growth factor receptor
FDA: Food and drug administration
GBM: Glioblastoma
GIST: Gastrointestinal stroma tumor
GVH: Graft versus host disease
HCC: Hepatocellular carcinoma
HL: Hemophagocytic lymphhistiocytosis
MB: Medulloblastoma
MDS: Myelodysplastic syndrome
MGMT: Methylguaninmethyltransferase
MPN: Myeloproliferative neoplasia
NC: Nasopharyngeal carcinoma
NTRK: Neurotrophic tyrosine kinase
PNH: Paroxysmal nightly heomglobinuria
SCD: Sickle cell disease
VEGF: Vasoendothelial growth factor

## Acknowledgements

We kindly thank Dr. Hanna Seidling (Cooperation Unit Clinical Pharmacology, Heidelberg) for help with the classification of designated compounds

